# A simple and quick PCR based method for detection of Omicron variant of SARS-CoV-2

**DOI:** 10.1101/2021.12.20.21268053

**Authors:** Apurvasinh Puvar, Ramesh Pandit, Armi M Chaudhari, Tasnim Travadi, Nitin Shukla, Chaitanya Joshi, Madhvi Joshi

## Abstract

SARS-CoV-2 pandemic has changed the global landscape since last two years. Against many challenges posed by the COVID-19 pandemic to the humanity, the pace of solutions created by mankind is exemplary; diagnostics, vaccines, alternate therapies, to name a few. With a rapidly changing virus strain, its early identification in the community can be a quick solution to trace the individuals and thus control its spread. This paper describes PCR based quick method for differentiation of Omicron variant of SARS-CoV-2 from other variants. Timely identification of this new variant will enable better management of pandemic control in the population.

**Graphical Abstract:** 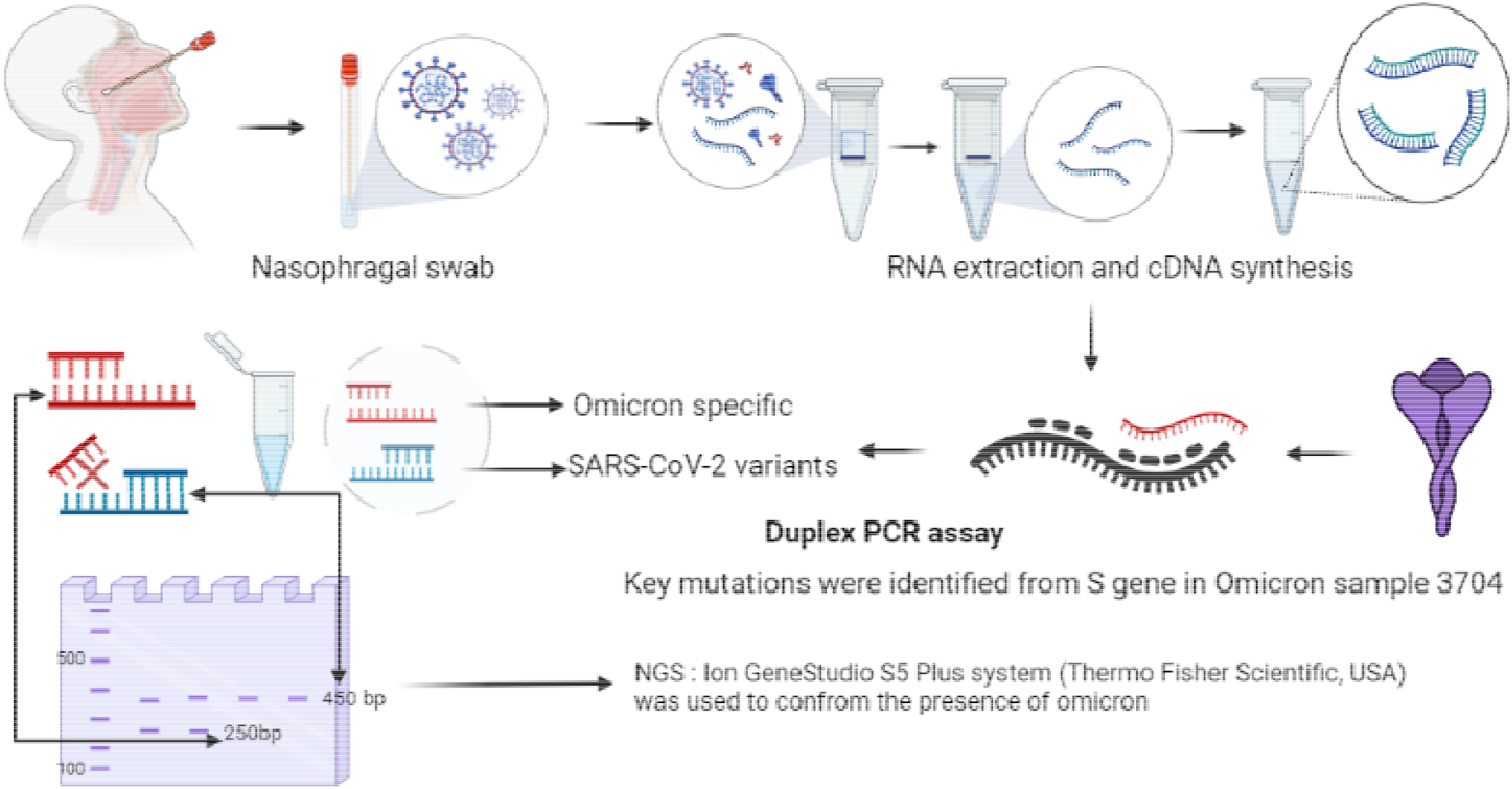

## Introduction

Omicron, B.1.1.529, (BA.1; according the new classification), the new emerging variant of concern (VOC) of SARS-CoV-2 has re-alerted the world again to the COVID-19 pandemic situation (1). Since the first case in Wuhan, China, scientists have discovered over 1500 SARS-CoV-2 lineages. Many of these lineages, however, have overpowered pandemic situations, such as the second wave in India caused by the Delta variant (B.1.167.2) of SARS-CoV-2 (2,3). This hyper mutated variant contains 15 mutations in RBD (receptor binding domain), leading to reduced binding of neutralizing antibodies but not complete escape from immunity (4). A second wave of the SARS-CoV-2 pandemic was sparked by the emergence of a delta variant with substantial mutations in the NTD (N-terminal domain), resulting in a prominent case of immunological escape (5,6). Omicron with concerning mutations in NTD, along with RBD might be more infectious and may lead to increased transmission as well as re-infection of SARS-CoV-2 (7). For better management of the pandemic situation, quick and reliable method to track the Omicron variant from the positive COVID-19 cases is very essential. Several laboratories have revealed that this VOC due to several spike mutations gives target failure/reduction in s gene based diagnostics, However, the SGTF can not be surely attributed to lineage Omicron. (https://www.who.int/news/item/26-11-2021-classification-of-omicron-(B.1.1.529)-sars-cov-2-variant-of-concern). Hence, at our laboratory, we developed a set of primers targeting the key mutations including insertions and deletions in spike protein encoding gene of this variant, allowing us to distinguish Omicron form the other SARS-CoV-2 lineages. This rapid PCR based test will help in early identification of Omicron cases from community and will help in better management in fight against COVID-19.

## 1. Materials & methods

Nasopharyngeal swab samples which we use in this study were collected by the Indian Council of Medical Research (ICMR, India) approved COVID-19 regional diagnosis laboratories (private and government). GBRC is also an authorized COVID-19 testing centre approved by Indian Council of Medical (ICMR) and also a member of Indian SARS-COV-2 Genome Sequencing Consortium (INSACOG), Government of India. Further, the work of SARS-CoV-2 genome sequencing at GBRC is approved the Institutional Ethical Committee of Gujarat Biotechnology Research Centre (GBRC), reference No. GBRC/Ethics/03/COVID19genome/2020-21. The COVID-19 positive samples which were sent by different laboratories for the SARS-CoV-2 genome sequencing were used in this study. Lowest RT-PCR ct values of these sample ranging from 16-33. QIAamp Viral RNA Mini Kit (Qiagen, Germany) was used to isolate viral RNA according to the manufacturer’s instructions (8). Later on, we used NGS Reverse Transcription Kit^®^ (Thermo Fisher Scientific, USA) to prepare complementary DNA (cDNA) (9). SARS-CoV-2 lineage in all the samples were confirmed by whole genome sequencing using Ion Ampliseq SARS-CoV-2 Research Panel and Ion GeneStudio S5 Plus system (Thermo Fisher Scientific, USA). To develop a PCR assay, primers were designed targeting the specific regions of the B.1.1.529 (BA.1) lineage of SARS-CoV-2. For the internal control, primers were designed from the conserved region of SARS-CoV-2 irrespective of the lineage. The sequences deposited in GISAID were considered for primer designing. For PCR assay, we used the same cDNA which was used for the genome sequencing. Experiments were divided into two groups, one using a Singleplex PCR test and the other using a Duplex PCR assay. PCR reaction mixture was prepared as shown in Table 1. Thermal Cycler conditions were set to 95°C for 3 minutes followed by 30 cycles of 95°C for 30 seconds, 55°C for 30 seconds and 72°C for 1 minute followed by final extension at 72°C for 5 minutes. Ten microliter of PCR products were run on 2% Agarose gel prepared in 0.5X TBE buffer at 120V for 15 min and visualized under UV light in the Biorad GelDoc® system.

**Table 1:**
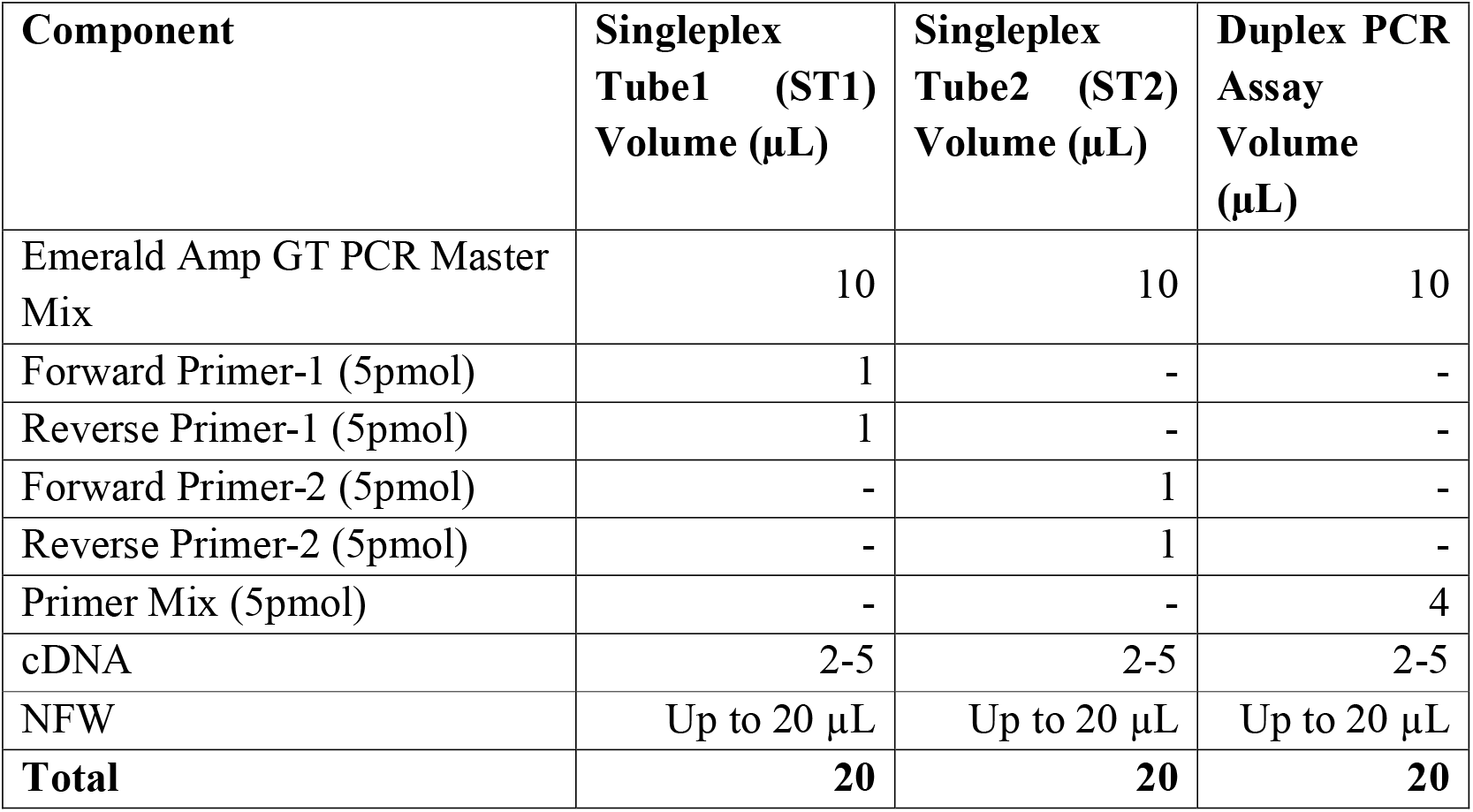
PCR reaction mixture for the Singleplex and Duplex PCR assay.

| **Component** | **Singleplex**<br>**Tube1 (ST1)**<br>**Volume (μL)** | **Singleplex**<br>**Tube2 (ST2)**<br>**Volume (μL)** | **Duplex PCR**<br>**Assay**<br>**Volume**<br>**(μL)** |
| --- | --- | --- | --- |
| Emerald Amp GT PCR Master Mix | 10 | 10 | 10 |
| Forward Primer-1 (5pmol) | 1 | - | - |
| Reverse Primer-1 (5pmol) | 1 | - | - |
| Forward Primer-2 (5pmol) | - | 1 | - |
| Reverse Primer-2 (5pmol) | - | 1 | - |
| Primer Mix (5pmol) | - | - | 4 |
| cDNA | 2-5 | 2-5 | 2-5 |
| NFW | Up to 20 μL | Up to 20 μL | Up to 20 μL |
| **Total** | **20** | **20** | **20** |

## 2. Results

Sample 3704 was the first case of Omicron detected in genome sequencing at our laboratory. Based on the genome sequence this and other Omicron sequences submitted in to GISAID database, we develop the primers. To begin, known omicron and other variants samples were utilised to validate the primer set developed. First, we use both primer sets from all of the samples to independently amplify the target DNA sequence (Figure 1). The upper lane in Figure 1 contains Omicron positive known sample 3704, which was Omicron positive and tested in duplicates. Later on, an unknown sample 4115, 4116, 4334 was discovered to be omicron positive using the PCR assay. Using whole genome sequencing also the samples were confirmed as a ‘Omicron’ variant, i.e. B.1.1.529 lineage of SARS-CoV-2, confirming the robustness of primes. The lineage in the other samples was B.1.617.2 (Delta) and other sub-lineages of the delta variant. Internal control (IC) band (450bp) was found in 11 out of 14 samples. The lack of amplification in IC in four samples could be due to a low viral load or a PCR reaction failure. In summary, four samples which are showing Omicron-specific bands (250bp), which were also validated by the genome sequencing. A band of internal control was also observed in these four samples. A non-specific band (950bp) was also found in some of the samples. However, while interpreting the results, this band may be overlooked.

**Figure 1:**
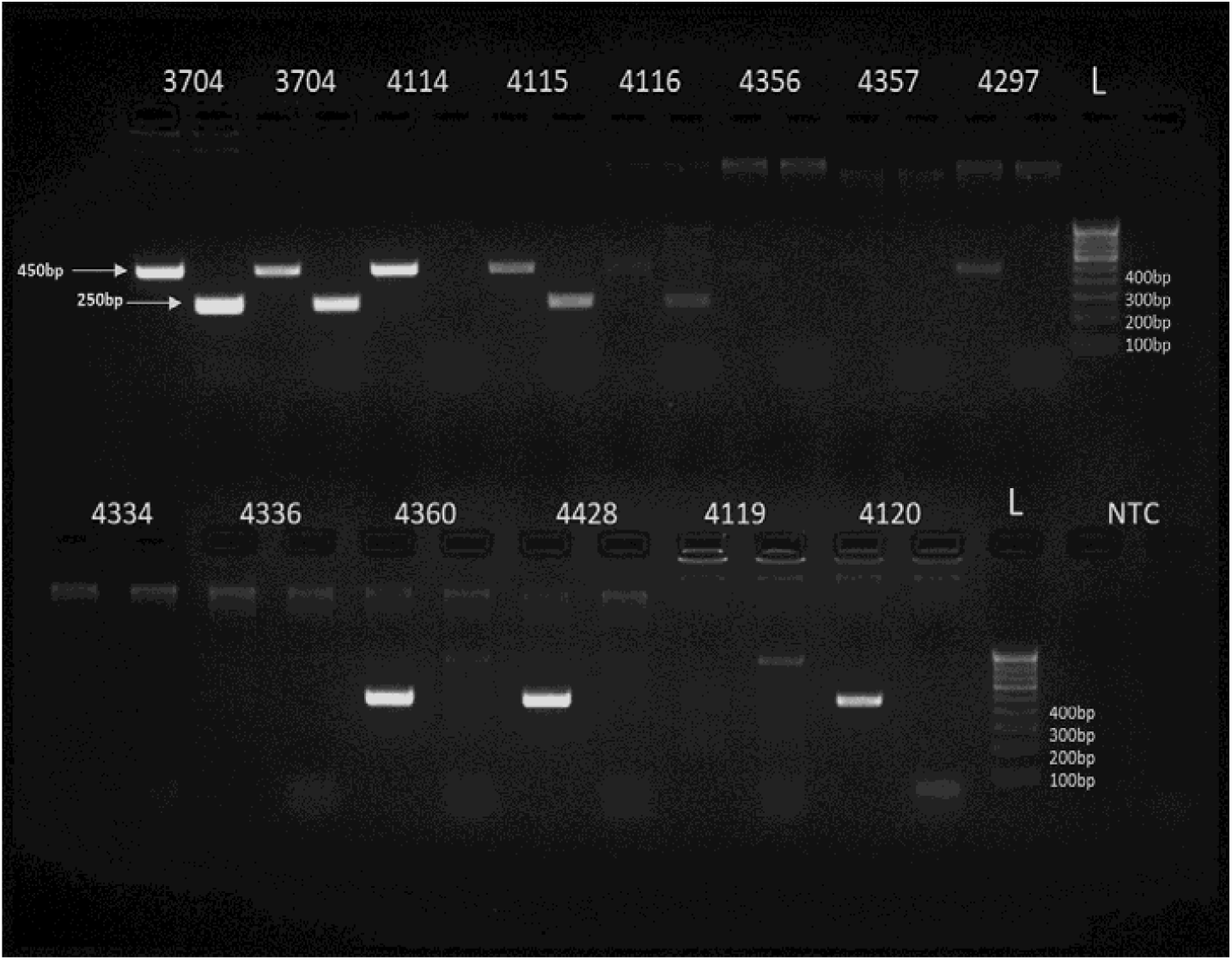
**Simplex** PCR based assay for the detection of Omicron (B.1.519.2): Primer set 1 resulted in to 250bp bands specific to Omicron. Primer set 2 resulted in to 450bp bands specific all SARS-CoV-2 variants. Numbering on well represents sample numbers for example. L is annoted for ladder (100bp). NTC is non-templet control. Bands other than 450bp and/or 250bp band may be ignored as, it may be nonspecific amplification.

Later, all four primers, that is internal control and Omicron specific were combined in a Duplex PCR assay reaction. We used two Omicron and one Delta lineage samples for the duplex PCR assay. In samples of the Omicron lineage, two bands of 450bp and 250bp were obtained as expected, whereas, only a single band of 450bp was amplified in other SARS-CoV-2 lineages (**Figure 2**). Table 2 describe how to interpret the results of a Duplex assay.

**Table 2:** Interpretation of Duplex PCR assay.

| **~450bp**<br>**Band (IC)** | **~250bp Band**<br>**(Omicron specific)** | **Interpretation** |
| --- | --- | --- |
| + | + | Omicron type of SARS-CoV-2 |
| + | - | SARS-CoV-2 (Other than Omicron) |
| - | + | Further confirmation required by sequencing |
| - | - | Less viral RNA load or failure of PCR reaction |

**Figure 2:**
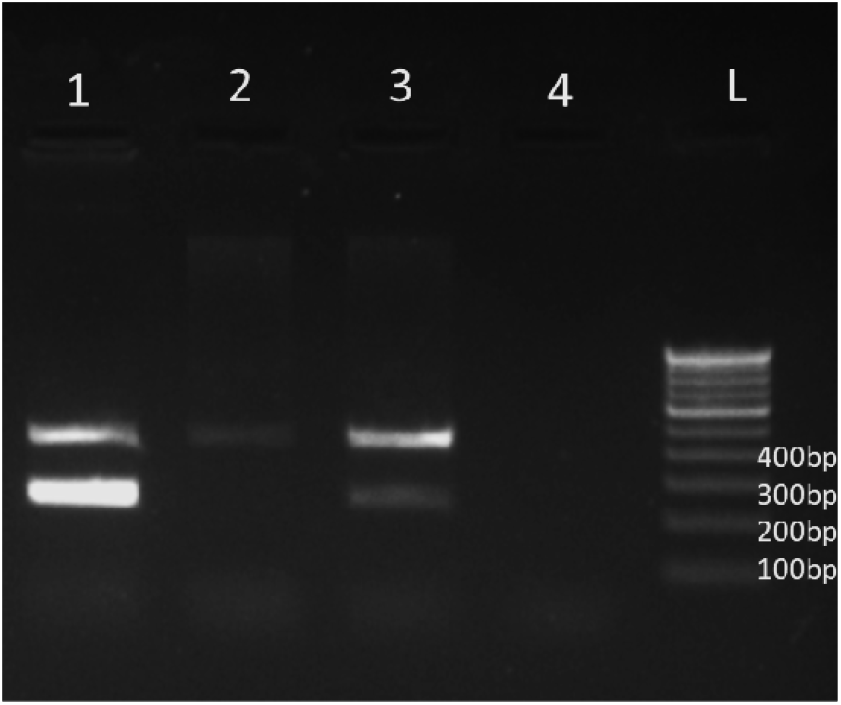
Duplex PCR assay results distinguishing Omicron from other SARS-CoV-2 variants on an agarose gel electrophoresis. Lane 1 & 3: presence of 250 bp band conform the presence of Omicron variant of SARS-CoV-2 in the infected patient, Lane 2: absence of 250bp band confirms other SARS-CoV-2 variant, Lane 5: NTC (Non-templet control). L: Ladder (100bp).

## 3. Discussion

Due to rapid community transmission, the SARS-CoV-2 pandemic had a significant impact on human health and the economy. Present SARS-CoV-2 pandemic diagnostic methods rely on antibody-antigen interactions, nucleic acid; however, viral nucleic acid detection by RT–PCR remains the cornerstone (10). In this pandemic, early detection of emerging SARS-CoV-2 lineages is essential for a synchronized worldwide response. We created a robust PCR-based detection kit that can distinguish between omicron from other SARS-CoV-2 variants. The Duplex PCR test reduced the cost of reagents and provide accurate results for the Omicron or no-Omicron SARS-CoV-2 variants. In sample having infection with Omicron, a 250-bp specific band will amplify, however in other variants; 250-bp amplification will be lacking due to primer sequence specificity. This type of quick PCR detection kit will be useful in overcoming the early detection and in turn will help to control the spread of Omicron variant.

## Data Availability

All data produced in the present study are available upon reasonable request to the authors

## 4. Data sharing

All data generated in these studies are available upon request made to the corresponding author, including primer sequences.

## 6. Acknowledge

Authors thanks Department of Science and Technology and Health and Family Welfare, (DST), Government of Gujarat for providing funding and support, respectively to carry out this work.

